# Assessment of machine learning algorithms in national data to classify the risk of self-harm among young adults in hospital: a retrospective study

**DOI:** 10.1101/2022.08.08.22278554

**Authors:** Anmol Arora, Louis Bojko, Santosh Kumar, Joseph Lillington, Sukhmeet Panesar, Bruno Petrungaro

**Author notes:** Correspondence to: Dr Anmol Arora, Trinity Hall, Trinity Lane, Cambridge CB2 1TJ, UK.

## Abstract

**Background:** Self-harm is one of the most common presentations at accident and emergency departments in the UK and is a strong predictor of suicide risk. The UK Government has prioritised identifying risk factors and developing preventative strategies for self-harm. Machine learning offers a potential method to identify complex patterns with predictive value for the risk of self-harm.

**Methods:** National data in the UK Mental Health Services Data Set were isolated for patients aged 18‒30 years who started a mental health hospital admission between Aug 1, 2020 and Aug 1, 2021, and had been discharged by Jan 1, 2022. Data were obtained on age group, gender, ethnicity, employment status, marital status, accommodation status and source of admission to hospital and used to construct seven machine learning models that were used individually and as an ensemble to predict hospital stays that would be associated with a risk of self-harm.

**Outcomes:** The training dataset included 23 808 items (including 1081 episodes of self-harm) and the testing dataset 5951 items (including 270 episodes of self-harm). The best performing algorithms were the random forest model (AUC-ROC 0.70, 95%CI:0.66-0.74) and the ensemble model (AUC-ROC 0.77 95%CI:0.75-0.79).

**Interpretation:** Machine learning algorithms could predict hospital stays with a high risk of self-harm based on readily available data that are routinely collected by health providers and recorded in the Mental Health Services Data Set. The findings should be validated externally with other real-world data.

**Funding:** This study was supported by the Midlands and Lancashire Commissioning Support Unit.

**Research in context:** 

**Evidence before this study:** Despite self-harm being repeatedly labelled as a national priority for psychiatric healthcare research, it remains challenging for clinicians to stratify the risk of self-harm in patients. National guidelines have highlighted deficiencies in care and attention is being paid towards the use of large datasets to develop evidence-based risk stratification strategies. However, many of the tools so far developed rely upon elements of the patient’s clinical history, which requires well curated datasets at a population level and previous engagement with care services at an individual level. Reliance upon elements of a patient’s clinical history also risks biasing against patients with missing data or against hospitals where data is poorly recorded.

**Added value of this study:** In this study, we use commissioning data that is routinely collected in the United Kingdom by healthcare providers with each hospital admission. Of the variables that were available for analysis, recursive feature elimination optimised our variable selection to include only age group, source of hospital admission, gender, and employment status. Machine learning algorithms were able to predict hospital episodes in which patients self-harmed in the majority of cases using a national dataset. Random forest and ensemble machine learning methods were the best-performing models. Sensitivity and specificity at predicting self-harm occurrence were 0.756 and 0.596, respectively, for the random forest model and 0.703 and 0.730 for the ensemble model. To our knowledge, this is the first study of its kind and represents an advance in the prediction of inpatient self-harm by limiting the amount of information required to make predictions to that which would be near-universally available at the point of the admission, nationally.

**Implications of all the available evidence:** There is a role for machine learning to be used to stratify the risk of self-harm when patients are admitted to mental health facilities, using only commissioning data that is easily accessible at the point of care. External validation of these findings is required as whilst the algorithms were tested on a large sample of national data, there remains a need for prospective studies to assess the real-world application of such machine learning models.

## Introduction

Self-harm and suicide are recognised as two serious adverse outcomes in the context of psychiatric illness, with repeated self-harm acting as the single strongest risk factor for suicide.^1^ In the UK, the National Health Service (NHS) Long Term Plan has highlighted the importance of ensuring access to mental health support, care, and treatment are accessible to all.^2^ The UK National Institute for Health and Care Excellence (NICE) has estimated that over 200 000 annual hospital attendances are due to self-harm, and has advised that all professionals working in the health and social care system have a responsibility to support at-risk patients.^3^ They have previously noted the importance of assessing the risk of repeat self-harm events, based on the characteristics of the harm, the person, and the circumstances.^4^ Adolescents and young adults are well recognised as the age group most at risk of self-harm,^5^ but there is important interplay between many modifiable clinical, psychosocial, demographic, and environmental factors associated with risk of self-harm.^6^ Identifying such risk factors and vulnerable patients is important to enable targeted and cost-effective interventions for the prevention of mental health deterioration and self-harm.^7,8^

Compared with research in child and adolescent populations, relatively little has been done to characterise the epidemiology of self-harm during hospital stays among young adults, despite this being a common occurrence.^9^ Indeed, adolescents and young adults are well recognised as the age group most at-risk of self-harm.^5^ A systematic review found that most instruments available for assessing risks of self-harm and suicide are not supported by sufficient evidence of accuracy,^10^ and another study found that none is sufficient to assess self-harm and suicide risks accurately.^11^ The Historical Clinical Risk Management-20 instrument, originally intended to assess risk of violence, has become particularly widely used and is mandated for use in secure services for forensic patients. However, NICE advises against the use of this and other risk assessment tools and scales to predict future risk of self-harm and suicide.^12^ Runeson and colleagues have called for more robust studies that are large enough to draw age- and diagnosis-specific conclusions on predictive validity.^10^ Previous research has largely focussed on elucidating elements of patients’ psychiatric histories that may influence their risk of self-harm.^13^ For example, a machine learning analysis identified specific emotional and behavioural presentations over 10 years that were associated with increased risk of self-harm in adolescents.^14^ However, reliance on patient history risks the issue of bias due to missing data because some patients are unable or unwilling to provide a complete psychiatric history, or those who belong to demographic groups associated with health data poverty may be under-represented.^15^ Use of data that are routinely collected and widely recorded might, therefore, improve prediction of self-harm.

In this national retrospective study, we explored whether machine learning predictive models based on mental health clinical commissioning data collected from mental health service providers in England could lead to risk stratification for episodes of self-harm. To our knowledge, this is the first use of commissioning data to develop a machine learning predictive model for self-harm at a national level. By using national clinical commissioning data collected from mental health service providers in the United Kingdom (UK) we use data that is available to care providers at the point of care delivery and we also advance on previous research efforts by not restricting our data to single-centre or regional hospital systems, which have been limited by generalisability to wider populations. The Mental Health Services Data Set (MHSDS) provides comprehensive demographic information but does not contain clinical information that would require access to patient notes. The use of machine learning enables the uncovering of patterns that would be infeasible to program due to potentially complex interactions between independent variables.

## Methods

### Study design and patients

This was a cross-sectional retrospective machine-learning study. Eligible patients were adults aged 18‒30 years who started a mental-health-related inpatient hospital spell starting between Aug 1, 2020 and Aug 1, 2021. We isolated data on patients from the MHSDS in the National Commissioning Data Repository (NCDR). This dataset includes data from patients who are in contact with mental health services in hospitals, the community, and outpatient clinics. Submitting data to the MHSDS is mandatory for NHS mental health providers and optional for non-NHS providers. The data collected are used to inform provider payments through the Mental Health Currencies and Payments system (formerly Payment by Results), and may be used for various purposes, such as clinical audit, research, and service design. The NCDR Platform is a securely accessed database that enables access to national data flows to allow NHS analysts and clinical commissioning groups (CCGs) to inform care planning based on data-driven intelligence. The dataset we used for the study included comprehensive anonymised demographic information but did not contain clinical information that would require access to patients’ notes. We excluded patients younger than 18 years because of potential differences in care and reporting between paediatric and adult facilities, and those with admissions that were ongoing on Jan 1, 2022. Unique anonymous patient identifiers were removed from the data for analysis.

Permission was granted by NHS England Data Services for the secondary analysis of anonymised data in the MHSDS, accessed through the NCDR secure server, for the purposes of this project.

### Selection of variables

Demographic data for each admission and information about whether there was a recorded episode of self-harm associated with the hospital stay were extracted. A maximum of one episode of self-harm was considered per hospital episode. If a patient had multiple hospital stays, a unique entry was created for each stay. Age was collapsed into a categorical variable consisting of four age groups (18‒ 21, 22‒24, 25‒27, and 28‒30 years). For remaining variables, missing data were imputed using missForest (version 1.4), with four iterations, to allow a random forest to be trained on observed data values and predict missing values. This method is well suited to the mixed types of data used in this study.

Seven variables recognised to be associated with and having plausible causative mechanisms for self-harm were selected for analysis: *age group, self-reported gender, ethnicity, employment status, marital status, accommodation status, and source of admission to hospital*.^16–18^ The addition of primary diagnosis, diagnosis history, substance misuse, and number of hours worked per week was considered, but these variables were discarded due to high proportions of missing data and potential overlap with those already selected. The selected variables are routinely collected alongside mental health stays and are often readily accessible to clinicians. Recursive feature elimination by tenfold cross-validation with five repeats was used in the training dataset to identify the optimal combination of variables for the predictive models. Of the seven original variables, age group, source of hospital admission, gender, and employment status were used in the final models. Dummy variables were constructed by one-hot encoding for each value of the categorical variables, resulting in 13 predictive variables.^19^

### Data processing

The dataset was split into a training dataset (80%, n=23808) and testing dataset (20%, n=5951). All data were scaled to be on the interval between zero and one to prevent disproportionate importance being assigned to variables with larger ranges of values. The scaling transform learnt on the training data was applied to the test data. As the positive outcome variable of an episode of self-harm was expected to be of low prevalence, the upSample function in R (version 4.1.0) was used to increase the sample rate for self-harm events and minimise algorithmic bias towards the majority class of no self-harm. Thus, 22 727 positive cases and 22 727 negative cases were used for training the algorithms in the training dataset.

### Statistical analysis

Statistical analyses were done on May 14, 2022, in R (version 4.1.0) via a secure NHS remote desktop server.^20^ Software packages used beyond the default R packages are listed in the Appendix A.

Tenfold cross-validation was used to train models, with area under receiver operating characteristic curve (AUC-ROC) used as the optimisation metric. Seven individual models based on diverse were constructed with the intention to cover a wide range of high-performing classification models, without hyperparameter optimisation: generalised linear; Bayesian generalised linear regression; radial kernel support vector machine; linear kernel support vector machine; random forest; neural network; boosted generalised linear. An ensemble model was constructed using all models, and weighted averages of predictions were applied as follows: no weighting in the generalised linear model; weight 0.1 in the Bayesian generalised linear regression and linear kernel support vector machine models, and weight 0.2 in the radial kernel support vector machine, random forest, neural network, and boosted generalised linear models. The predictions of the random forest model were weighted relatively more than the other models due to its high accuracy, sensitivity, and specificity.

We compared accuracy, sensitivity, specificity, positive predictive value, negative predictive value, and AUC-ROC in each model. Using the glm function in R, a binomial regression model using the whole dataset was constructed to identify variables correlated with the outcome variable and calculate coefficients in order to assess the overall directionality of the relationships. Variables were tested for correlation with the view that highly correlated variables should be removed to limit the risk of harmful bias and increased variance. No variables were removed for this reason. We created a variable importance plot using the Caret Varimp() function for each model to investigate whether any factors were consistently associated with a high risk of self-harm episodes.

The STROBE checklist items were considered when reporting the findings of this study (Appendix B).^21^

## Results

### Descriptive statistics

There were a total of 228 826 hospital stays starting between August 1, 2020 and August 1, 2021 were reported in the MHSDS for patients aged between 18 to 30. Of these, 79 384 had discharge dates recorded before Jan 1, 2022. Duplicate entries were removed, with the most recent entry being retained. The total number of unique relevant hospital episodes was 29 759. Table 1 illustrates the demographics of the study population, categorised by age group. The prevalence of the outcome variable, an episode of self-harm, is also included. The number of self-harm events was 1351 (4.5%). The training dataset included 23 808 items and the testing dataset 5951 items, including 1081 and 270 episodes of self-harm respectively.

**Table 1:**
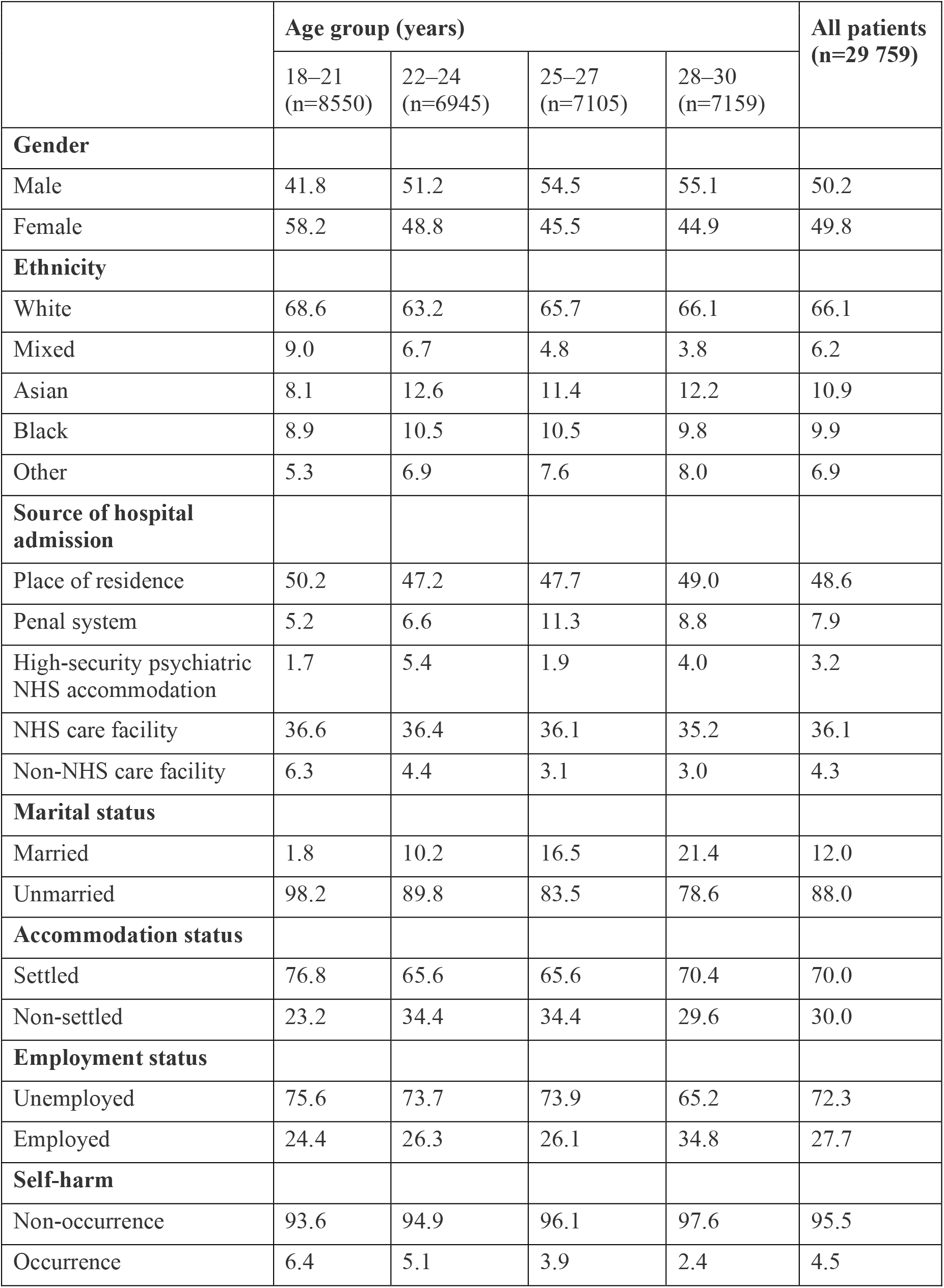
Characteristics of the study population. Data are %.

Variables were tested for correlation with the view that highly correlated variables should be removed to limit the risk of harmful bias and increased variance. Figure 1 illustrates a correlation matrix between the variables, including those that were not included in the final models. We found no highly correlated variables that needed removal, with the strongest correlation, between marital status and employment status, having a Pearson correlation coefficient of only 0.41 (Figure 1).

**Figure 1:**
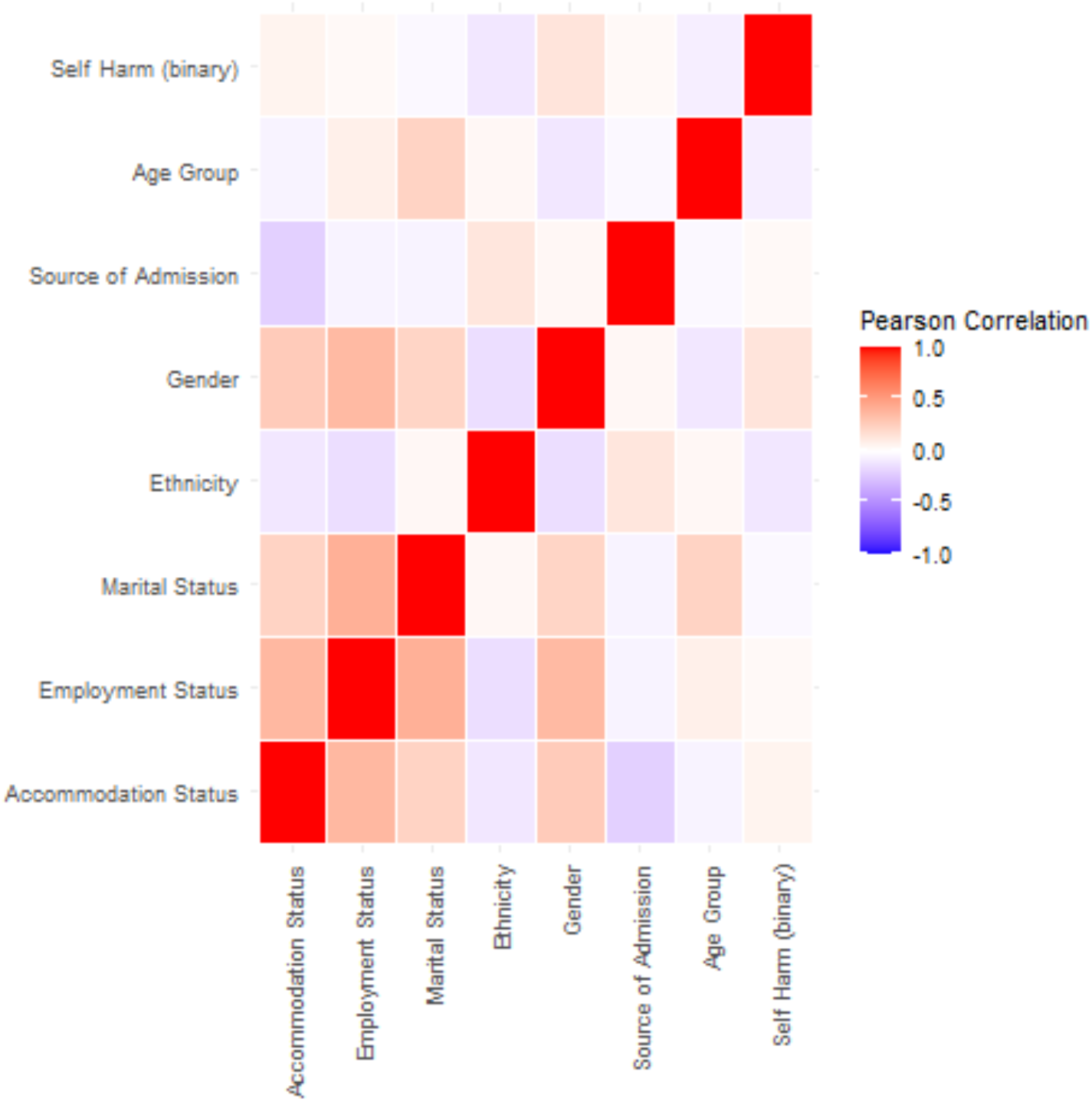
Correlation heatmap for variables used in the machine learning model. The strongest correlation was between employment status and marital status (coefficient 0.41), which did not meet the threshold for removal.

The results of the seven individual models and the ensemble model in the test dataset are shown in Table 2. The ensemble model appears to be the best performing model, both by accuracy and AUC-ROC, although it does have a relatively high number of false negatives.

**Table 2:** Summary performance measures of predictive models in the testing dataset. The test dataset included 5951 items. Data are presented with 95% confidence intervals. Random forest and ensemble models produced the highest AUC-ROC values. The ensemble model also produced the highest accuracy and specificity. AUC-ROC=area under receiver operating characteristic curve.

|  | Accuracy | Sensitivity | Specificity | True positives | True negatives | False positives | False negatives | AUC-ROC |
| --- | --- | --- | --- | --- | --- | --- | --- | --- |
| Generalised linear regression | 0.585 | 0.73 (0.67–0.78) | 0.58 (0.56–0.59) | 197 | 3283 | 2398 | 73 | 0.69 (0.65–0.73) |
| Bayesian generalised linear regression | 0.585 | 0.73 (0.67–0.78) | 0.58 (0.56–0.59) | 197 | 3283 | 2398 | 73 | 0.69 (0.65–0.73) |
| Radial kernel support vector machine | 0.606 | 0.73 (0.67–0.78) | 0.60 (0.59–0.61) | 197 | 3407 | 2274 | 73 | 0.68 (0.64–0.72) |
| Linear kernel support vector machine | 0.529 | 0.76 (0.70–0.80) | 0.52 (0.51–0.53) | 204 | 2944 | 2737 | 66 | 0.65 (0.61–0.69) |
| Neural network | 0.600 | 0.73 (0.67–0.78) | 0.59 (0.58–0.61) | 196 | 3374 | 2307 | 74 | 0.69 (0.65–0.73) |
| Random forest | 0.577 | 0.76 (0.70–0.80) | 0.57 (0.56–0.58) | 204 | 3231 | 2450 | 66 | 0.7 (0.66–0.74) |
| Boosted generalised linear regression | 0.562 | 0.75 (0.69–0.80) | 0.55 (0.54–0.57) | 202 | 3141 | 2540 | 68 | 0.69 (0.65–0.73) |
| Ensemble | 0.705 | 0.70 (0.69–0.72) | 0.73 (0.68–0.78) | 203 | 4047 | 1706 | 75 | 0.77 (0.75–0.79) |

Figure 2 compares the results of each model based on receiver operating characteristic (ROC) curves derived from the performance of the algorithms on the test dataset.

**Figure 2:**
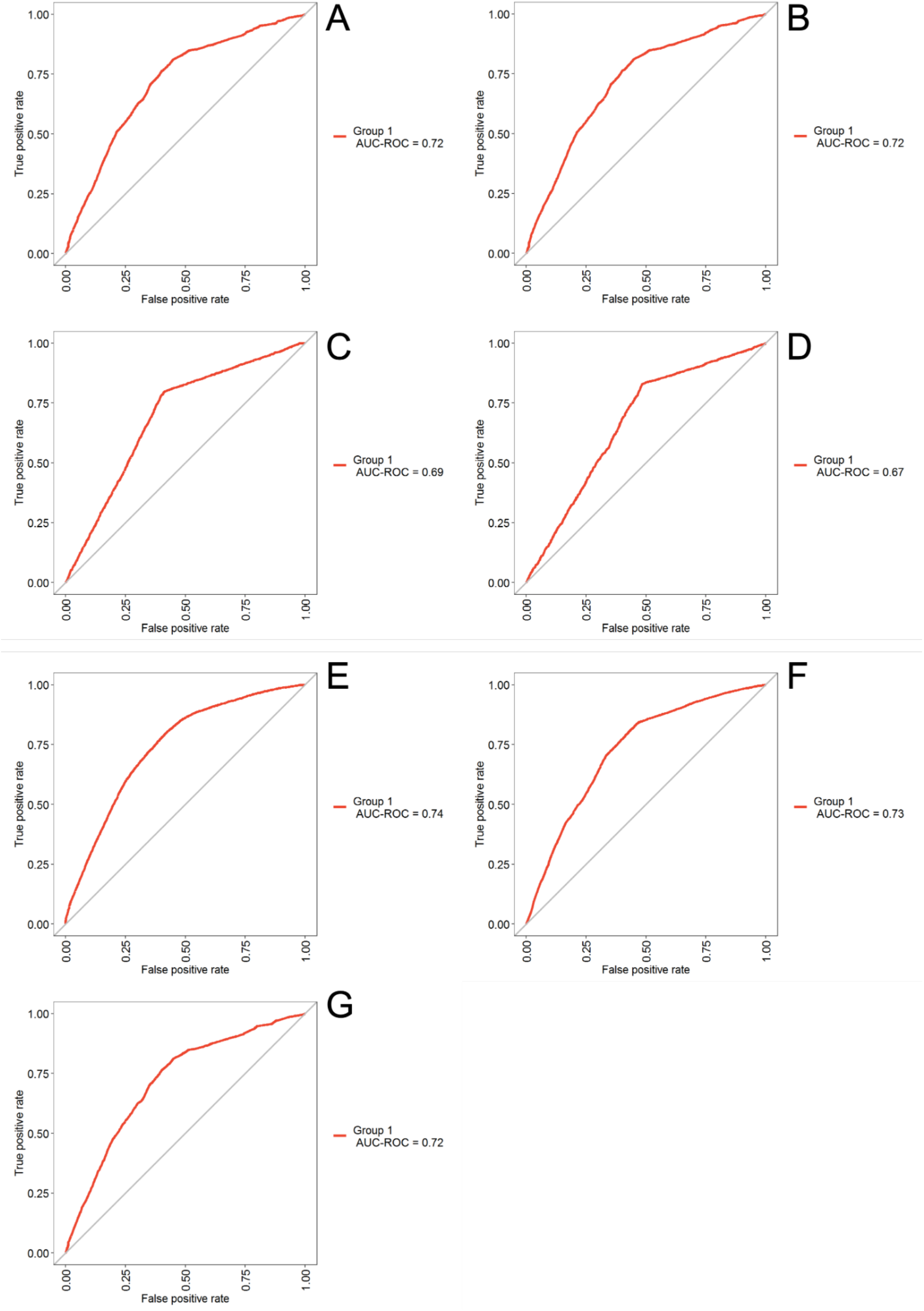
Receiver operating characteristic (ROC) curves for the individual predictive models, based on the training dataset. The training dataset included 23 808 items. (A) Generalised linear regression model. (B) Bayesian generalised linear regression model. (C) Radial kernel support vector machine model. (D) Linear kernel support vector machine model. (E) Neural network model. (F) Random forest model. (G) Boosted generalised linear regression model. High performing models tend to occupy the top left of the plots with poor models lying on or below the 45-degree diagonal of the ROC space. AUC-ROC=area under receiver operating characteristic curve.

### Variable importance

The variable importance plots for each model are shown in Figure 3. The plot does not specify whether the variable was a positive or negative predictor but the directionality of the relationship can be suggested by multiple regression. Multiple regression analysis showed that variables associated with an increased risk of self-harm were: female, source of admission anywhere except residence, unsettled accommodation, white ethnicity, younger age and unemployment. (Appendix C).

**Figure 3:**
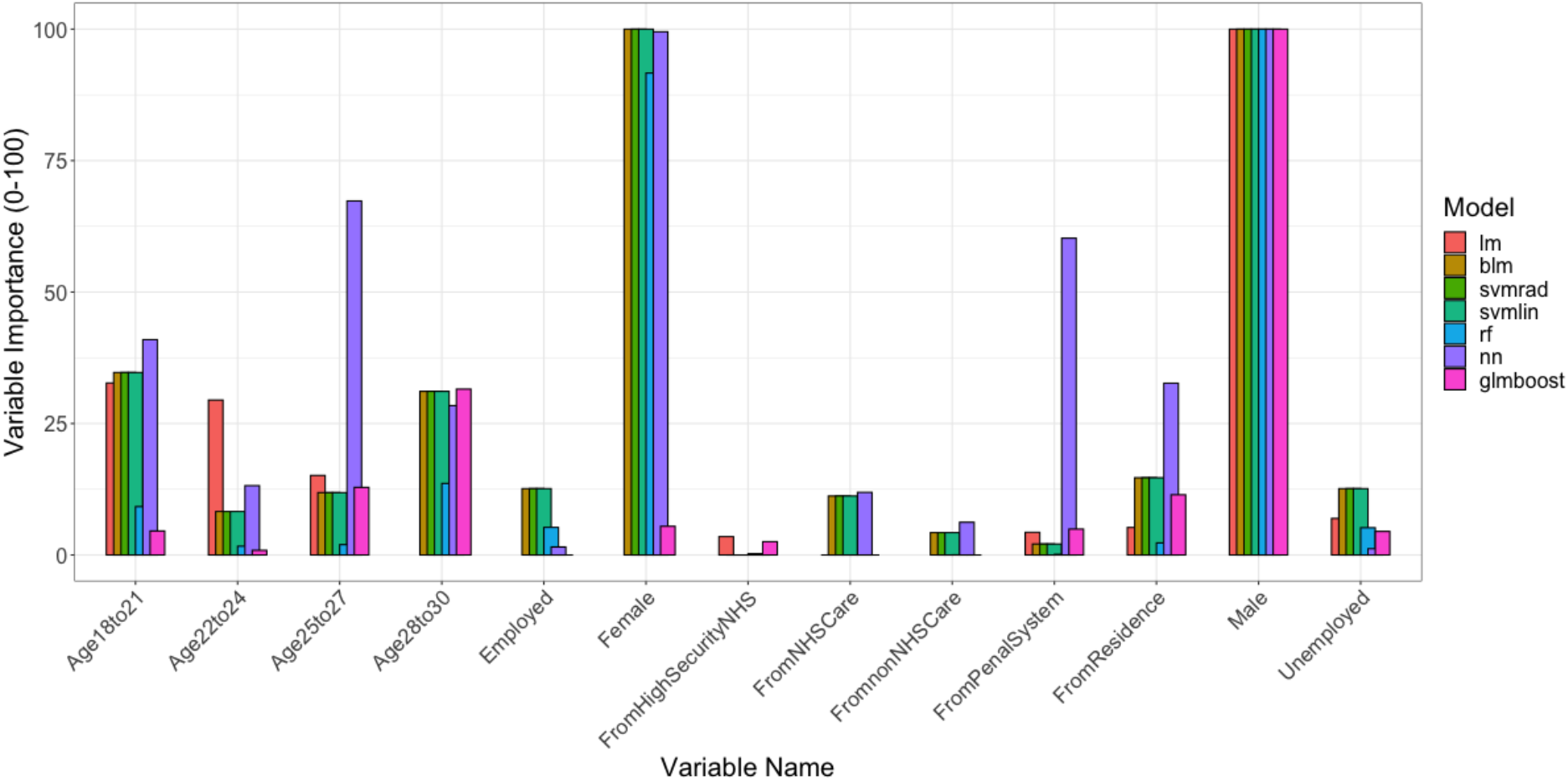
Variable importance plots by model. Variables with importance value 0 are not visible. lm=generalised linear regression model; blm=Bayesian generalised linear regression model; svmrad=radial kernel support vector machine model; svmlin=linear kernel support vector machine model; nn=neural network model; rf=random forest model; glmboost=boosted generalised linear regression model.

## Discussion

### Summary of key findings

With commissioning data that would be available at the point of care, we found that various machine learning algorithms could identify complex patterns with predictive value for the risk of self-harm among adults in hospital. The best performing model correctly predicted 70.5% of individuals who self-harmed during a hospital episode (sensitivity), with a specificity of 73%. Studying variable importance identified that age, gender, employment status and source of admission could potentially be used in clinical care to determine the risk of self-harm. Our findings add to the growing body of evidence that machine learning may have operational benefits in mental health care that can contribute to meeting the NHS’s national priorities.

All the algorithms tested achieved a respectable level of sensitivity and specificity. Sensitivity might be more relevant than specificity in management of self-harm, as the risks of providing extra intervention are unlikely to compare to those associated with not intervening.

### Comparison to existing literature

Several attempts have been made to develop predictive models for self-harm and suicide from large datasets, and a systematic review noted that machine learning has is likely to enhance the yield of accurate predictions compared to traditional statistical techniques.^22^ In the USA, an algorithm was developed to predict the risk of suicide and self-harm among women with depression, bipolar disorder, and chronic psychosis, and it yielded AUC-ROCs of 0.71‒0.73 with accuracy of 84%.^23^ An Australian study assessed machine learning algorithms for predicting self-harm among patients presenting to youth mental health services. The AUC-ROCs were 0.744–0.755 but the study was limited by sample size (n=1962 patients) and class imbalance (320 [16%] self-harm *vs* 1642 [84%] no self-harm).^24^

In this study we focused on the use of commissioning level data, but there has also been interest in using clinical text from electronic health-care records in the USA. Analysis of these records has yielded high accuracy but the approach is only applicable to hospitals that universally use electronic records, which is rare in the UK. Obeid et al investigated whether text processing of clinical notes could predict self-harm.^25^ Machine learning algorithms were trained on notes made over a maximum period of 90 days within 1‒6 months before the index event of self-harm. Predictive accuracy for the best-performing model in a test set of 200 patients was 79% (AUC-ROC 0.88). Higher levels of accuracy have been achieved in clinical text notes with techniques such as natural language processing, even without considering demographic information, but this approach requires accurate reporting of ICD diagnostic codes.^25^

Our models were trained on only a handful of readily available features. However, the application of natural language processing to structured electronic health records allows construction of hundreds of features. For example, one study used up to 2126 different features based on structured and unstructured data, including data from clinical notes, demographics, diagnoses, health-care use, and medication history, to predict suicide attempts.^26^ Among four models assessed, the best performance was achieved with 1726 variables, yielding AUC-ROCs of 0.919‒0.932. The value of using a patient’s most recent data rather than long-term historical data was shown in an American study which found that accuracy of predictions increased from AUC-ROCs 0.75‒0.76 when based on data from the previous 720 days to 0.82‒0.85 when using data from within 7 days of the event.^27^

Risk factors for repetition of self-harm have been widely studied and are varied, with longstanding psychosocial vulnerabilities having the most consistent evidence.^28^ However, identifying these risk factors requires access to the patient’s psychiatric history, which is recorded in different data formats by different hospitals. The results of our study suggest that it may be possible to use widely available commissioning data as a high-level analysis of which patients would benefit from a more in-depth assessment of personal risk factors from their psychiatric history at a local level.

### Implications of findings

Our study relied on commissioning data that would generally be available at the time of hospital admission. By contrast, many previous studies have used elements of clinical history, but these may be less readily available or unreliably reported at the point of admission. Additionally, clinicians’ judgement of patients’ risk of self-harm is unreliable.^29^ Furthermore, while locally developed risk tools and scales, such as the SAD PERSONS risk scale, can be used to assess risk, their accuracy is not well validated.^10^ Without this supporting evidence, there is little consensus over the most suitable tools.^30^ Our findings, therefore, could be a step towards filling an important research gap. Provision of care for patients presenting with self-harm remains variable despite refreshed national guidance.^31^ Another limitation of previous research is the restriction to single or regional hospital systems, which reduces generalisability to other providers, who may record patient data in a different format. Our study overcomes this by using a dataset which is populated by all mental health providers nationally as part of routine care delivery.

The results of this study could in the future be used to inform local and public health measures to identify and support patients who are at high risk of self-harm. At the time of admission, patients could be identified as being high-risk of self-harm and further risk-assessment performed or appropriate interventions prepared. Methods of reducing self-harm in vulnerable patients once they have been identified have been well explored, including by implementing protective strategies or engaging with pharmacological or psychological treatments.^32,33^ The evidence taken together, therefore, could lead to effective treatment mechanisms to prevent self-harm in at-risk individuals being put in place.

Self-harm is one of the most common presentations in accident and emergency departments in the UK, and in England alone hospital management of self-harm costs an estimated £162 million per year.^34,35^ The prioritisation of suicide and self-harm prevention by the UK government indicates that there remains a strong health economic argument for targeted strategies. Public Health England has recognised the role of using national datasets to identify high-risk groups.^36^ This study benefits from the use of national data, meaning that the findings are likely to be generalisable to inpatients in the UK. The proportion of inpatients self-harming was broadly consistent with previous rates, which are recognised as rising.^37^ Additionally, while the issue of health data poverty is known to affect machine learning research due to under-representation of some subgroups of patients, our dataset showed considerable diversity. Across the UK only approximately 14% of the population belongs to ethnic minority groups, such patients accounted for roughly 34% of our dataset.^38^ This difference is explained by the over-representation of ethnic minority patients amongst those in the MHSDS.

### Limitations

This study was limited by the dataset we used only capturing episodes of self-harm associated with inpatient treatment in mental health facilities. Therefore, the findings may not be generalisable to a community. We recorded a maximum of one self-harm event per hospital stay to limit algorithmic bias and, although we aimed to use the latest sociodemographic information for each patient, there may have been instances where this information was incorrect, out of date, or missing. However, no variables with disproportionate amounts of missing data (>50%) were included in the models and multiple imputation was used to account for missing data for included variables. Machine learning as a statistical technique is limited by generalisability and the risk of overfitting.^39^ Our findings are relatively resistant to these limitations compared to previous studies because of the large national dataset used, but external validation would be required to establish reproducibility in a real-world setting. We were unable to assess potential causative mechanisms for the risk of self-harm. As the data is only collected when patients present to mental health care services, it was not possible to determine whether recent changes in the patient’s information were responsible for self-harm risks, for example if a patient recently became homeless or unemployed. The upper age cut-off for defining young adults is debatable and this study chose a limit of 30 years in recognition of the fact that the boundary could range from 24 to 35 years.^40^ Studying variable importance and performing multiple regression to determine whether variables were positive or negative predictors of self-harm was used to attempt to overcome the black-box phenomenon, but that assumes the directionality of the relationship used by the algorithm is concordant with the directionality found by multiple regression.

### Future directions of research

This study progresses the use of statistical analysis by machine learning to inform targeted interventions to reduce self-harm and suicide in the UK, in line with national health priorities. To our knowledge, this is the first study to utilise national data towards the prediction of inpatient self-harm events using commissioning data that would be available at the point of care. In this way, our study addresses two substantial limitations plaguing previous research efforts: regional generalisability by using a national dataset and clinical system generalisability by using universally recorded commissioning data. This work may be used as a benchmark for future modelling studies of this nature, as machine learning methods evolve. Future avenues of research should explore, with prospective studies, whether attempts to intervene and contact patients who are suggested as being at increased risk by machine learning algorithms can reduce the numbers of events. The use of commissioning data limited our analysis to episodes of self-harm that were associated with hospital stays. Further research might also explore the application of this analysis to similar commissioning data from digital primary care records, in order to target self-harm beyond the hospital setting.

## Data Availability

Information about the Mental Health Services Data Set is available at https://digital.nhs.uk/data-and-information/data-collections-and-data-sets/data-sets/mental-health-services-data-set/. The dataset documentation is publicly accessible but the data is only available through England NHS Data Services for approved uses.

## Acknowledgments

This study was supported by the Midlands and Lancashire Commissioning Support Unit. We thank Rachel Ashton for editorial assistance.

## APPENDIX

### APPENDIX A Additional software packages used in the statistical analysis

Additional software packages used in the statistical analysis
| Package Name | Version Number | Package Documentation | Date last accessed |
| --- | --- | --- | --- |
| ggplot2 | 3.3.3 | https://cran.r-project.org/web/packages/ggplot2/ | 14th May 2022 |
| reshape2 | 1.4.4 | https://cran.r-project.org/web/packages/reshape2/ | 14th May 2022 |
| odbc | 1.3.2 | https://cran.r-project.org/web/packages/odbc/ | 14th May 2022 |
| DBI | 1.1.1 | https://cran.r-project.org/web/packages/DBI/ | 14th May 2022 |
| plyr | 1.8.6 | https://cran.r-project.org/web/packages/plyr/ | 14th May 2022 |
| dplyr | 1.0.6 | https://cran.r-project.org/web/packages/dplyr/ | 14th May 2022 |
| dbplyr | 2.1.1 | https://cran.r-project.org/web/packages/dbplyr/ | 14th May 2022 |
| tidyverse | 1.3.1 | https://cran.r-project.org/web/packages/tidyverse/ | 14th May 2022 |
| skimr | 2.1.3 | https://cran.r-project.org/web/packages/skimr/ | 14th May 2022 |
| missForest | 1.4 | https://cran.r-project.org/web/packages/missForest/ | 14th May 2022 |
| caret | 6.0.88 | https://cran.r-project.org/web/packages/caret/ | 14th May 2022 |
| psych | 2.1.3 | https://cran.r-project.org/web/packages/psych/ | 14th May 2022 |
| MLeval | 0.3 | https://cran.r-project.org/web/packages/MLeval/ | 14th May 2022 |

### APPENDIX B STROBE Statement—checklist of items that should be included in reports of observational studies

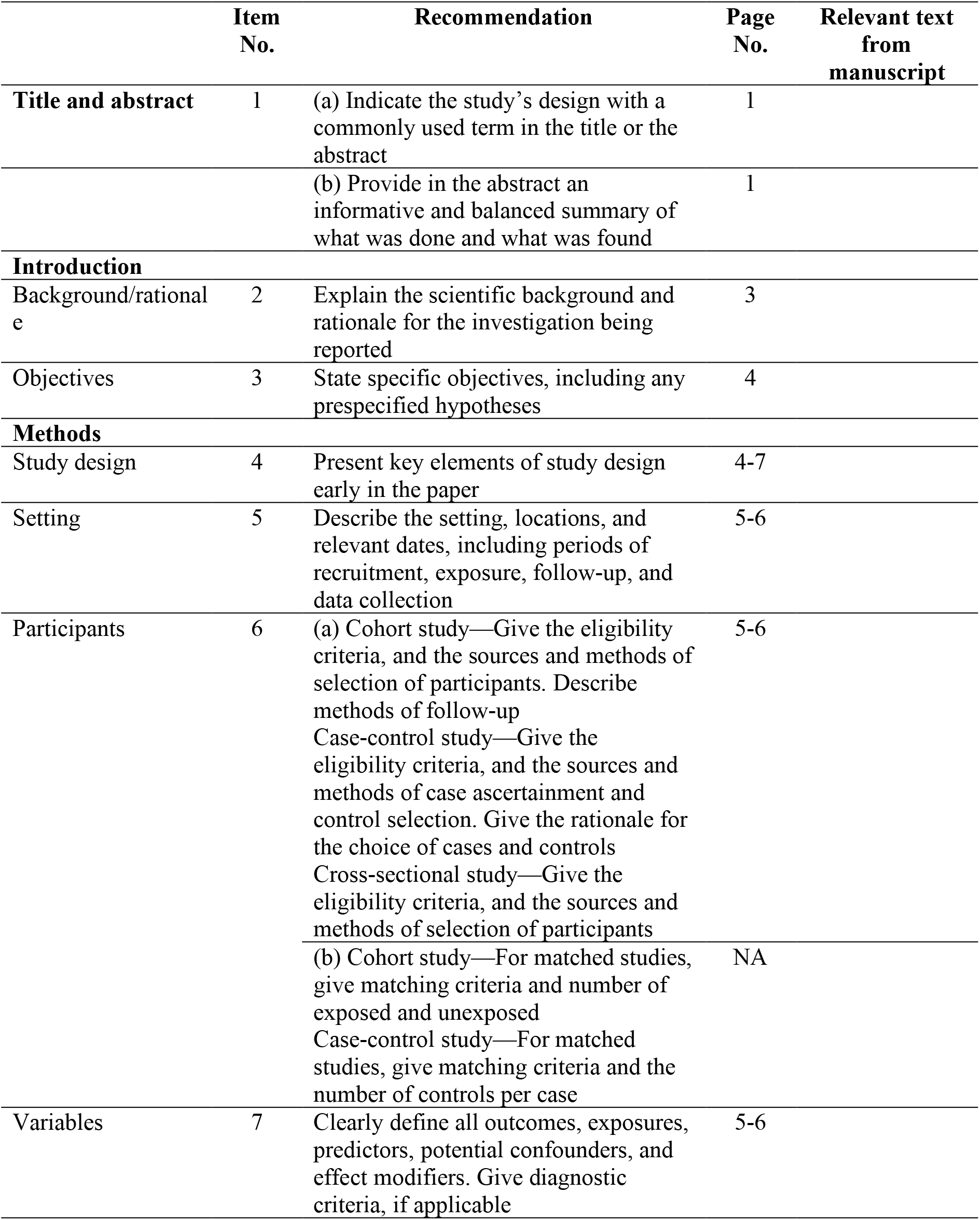
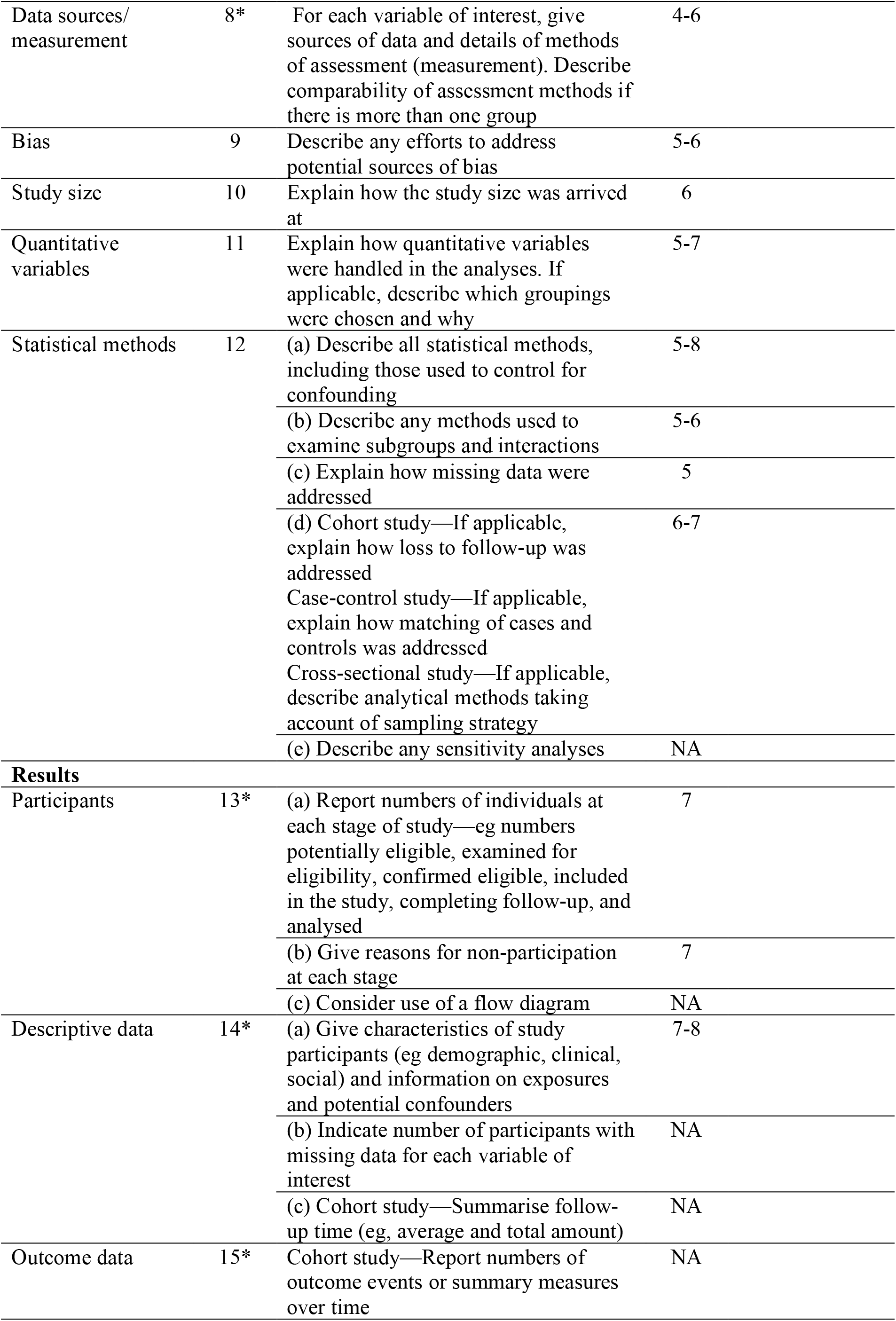
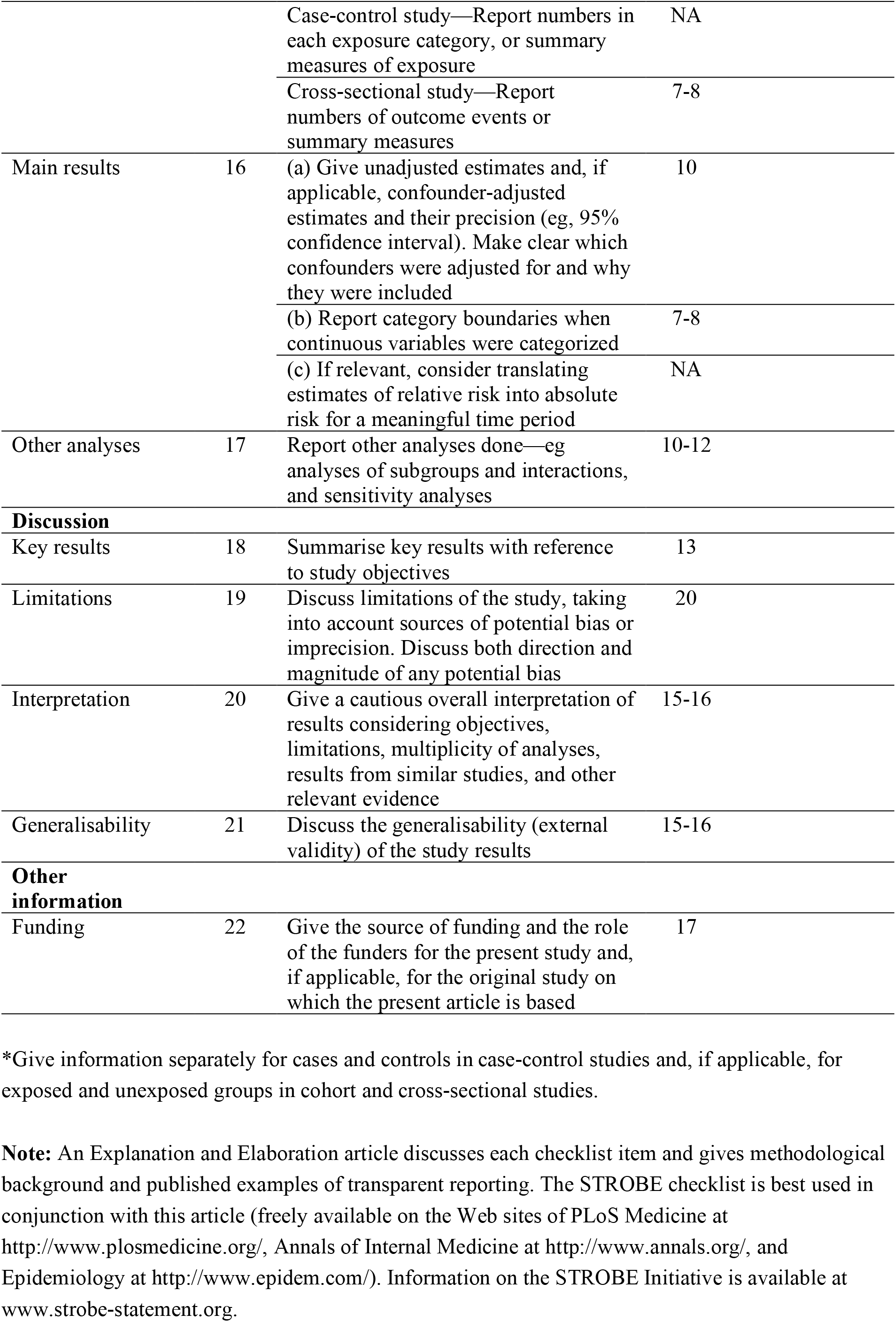

|  | Item No. | Recommendation | Page No. | Relevant text from manuscript |
| --- | --- | --- | --- | --- |
| Title and abstract | 1 | (a) Indicate the study’s design with a commonly used term in the title or the abstract | 1 |  |
|  |  | (b) Provide in the abstract an informative and balanced summary of what was done and what was found | 1 |  |
| Introduction |  |  |  |  |
| Background/rationale | 2 | Explain the scientific background and rationale for the investigation being reported | 3 |  |
| Objectives | 3 | State specific objectives, including any prespecified hypotheses | 4 |  |
| Methods |  |  |  |  |
| Study design | 4 | Present key elements of study design early in the paper | 4-7 |  |
| Setting | 5 | Describe the setting, locations, and relevant dates, including periods of recruitment, exposure, follow-up, and data collection | 5-6 |  |
| Participants | 6 | (a) Cohort study—Give the eligibility criteria, and the sources and methods of selection of participants. Describe methods of follow-up<br>Case-control study—Give the eligibility criteria, and the sources and methods of case ascertainment and control selection. Give the rationale for the choice of cases and controls<br>Cross-sectional study—Give the eligibility criteria, and the sources and methods of selection of participants | 5-6 |  |
|  |  | (b) Cohort study—For matched studies, give matching criteria and number of exposed and unexposed<br>Case-control study—For matched studies, give matching criteria and the number of controls per case | NA |  |
| Variables | 7 | Clearly define all outcomes, exposures, predictors, potential confounders, and effect modifiers. Give diagnostic criteria, if applicable | 5-6 |  |

|  |  |  |  |
| --- | --- | --- | --- |
| Data sources/<br>measurement | 8* | For each variable of interest, give sources of data and details of methods of assessment (measurement). Describe comparability of assessment methods if there is more than one group | 4-6 |
| Bias | 9 | Describe any efforts to address potential sources of bias | 5-6 |
| Study size | 10 | Explain how the study size was arrived at | 6 |
| Quantitative variables | 11 | Explain how quantitative variables were handled in the analyses. If applicable, describe which groupings were chosen and why | 5-7 |
| Statistical methods | 12 | (a) Describe all statistical methods, including those used to control for confounding | 5-8 |
|  |  | (b) Describe any methods used to examine subgroups and interactions | 5-6 |
|  |  | (c) Explain how missing data were addressed | 5 |
|  |  | (d) Cohort study—If applicable, explain how loss to follow-up was addressed<br>Case-control study—If applicable, explain how matching of cases and controls was addressed<br>Cross-sectional study—If applicable, describe analytical methods taking account of sampling strategy | 6-7 |
|  |  | (e) Describe any sensitivity analyses | NA |
| **Results** |  |  |  |
| Participants | 13* | (a) Report numbers of individuals at each stage of study—eg numbers potentially eligible, examined for eligibility, confirmed eligible, included in the study, completing follow-up, and analysed | 7 |
|  |  | (b) Give reasons for non-participation at each stage | 7 |
|  |  | (c) Consider use of a flow diagram | NA |
| Descriptive data | 14* | (a) Give characteristics of study participants (eg demographic, clinical, social) and information on exposures and potential confounders | 7-8 |
|  |  | (b) Indicate number of participants with missing data for each variable of interest | NA |
|  |  | (c) Cohort study—Summarise follow-up time (eg, average and total amount) | NA |
| Outcome data | 15* | Cohort study—Report numbers of outcome events or summary measures over time | NA |
|  |  | Case-control study—Report numbers in each exposure category, or summary measures of exposure | NA |
|  |  | Cross-sectional study—Report numbers of outcome events or summary measures | 7-8 |
| Main results | 16 | (a) Give unadjusted estimates and, if applicable, confounder-adjusted estimates and their precision (eg, 95% confidence interval). Make clear which confounders were adjusted for and why they were included | 10 |
|  |  | (b) Report category boundaries when continuous variables were categorized | 7-8 |
|  |  | (c) If relevant, consider translating estimates of relative risk into absolute risk for a meaningful time period | NA |
| Other analyses | 17 | Report other analyses done—eg analyses of subgroups and interactions, and sensitivity analyses | 10-12 |
| **Discussion** |  |  |  |
| Key results | 18 | Summarise key results with reference to study objectives | 13 |
| Limitations | 19 | Discuss limitations of the study, taking into account sources of potential bias or imprecision. Discuss both direction and magnitude of any potential bias | 20 |
| Interpretation | 20 | Give a cautious overall interpretation of results considering objectives, limitations, multiplicity of analyses, results from similar studies, and other relevant evidence | 15-16 |
| Generalisability | 21 | Discuss the generalisability (external validity) of the study results | 15-16 |
| **Other information** |  |  |  |
| Funding | 22 | Give the source of funding and the role of the funders for the present study and, if applicable, for the original study on which the present article is based | 17 |

\*Give information separately for cases and controls in case-control studies and, if applicable, for exposed and unexposed groups in cohort and cross-sectional studies.
**Note:** An Explanation and Elaboration article discusses each checklist item and gives methodological background and published examples of transparent reporting. The STROBE checklist is best used in conjunction with this article (freely available on the Web sites of PLoS Medicine at http://www.plosmedicine.org/, Annals of Internal Medicine at http://www.annals.org/, and Epidemiology at http://www.epidem.com/). Information on the STROBE Initiative is available at www.strobe-statement.org.

### APPENDIX C

**Table A1:** Results of a generalised linear regression model between predictive variables and self-harm. The reference categories are: white ethnicity, male gender, unemployed, unsettled accommodation, from residence, age 18 to 21.

| *Variable* | *Coefficient* | *Error* | *z value* | *Pr(>\|z\|)* |
| --- | --- | --- | --- | --- |
| *(Intercept)* | -3.93454 | 0.10221 | -38.067 | -3.934540.10221-38.067 |
| *Settled Accommodation* | 0.5063 | 0.08437 | 5.996 | 0.50630.084375.996 |
| *Employed* | -0.46999 | 0.06563 | -7.117 | -0.469990.06563-7.117 |
| *Mixed* | -1.29428 | 0.15621 | -7.853 | -1.294280.15621-7.853 |
| *Asian* | -1.36432 | 0.13617 | -8.679 | -1.364320.13617-8.679 |
| *Black* | -2.26732 | 0.22776 | -10.185 | -2.267320.22776-10.185 |
| *Other* | -0.91455 | 0.19315 | -5.255 | -0.914550.19315-5.255 |
| *Female* | 1.43583 | 0.07549 | 18.835 | 1.435830.0754918.835 |
| *From court, penal system, etc* | 0.57077 | 0.12393 | 4.655 | 0.570770.123934.655 |
| *From another NHS provider* | 0.59224 | 0.1608 | 3.784 | 0.592240.16083.784 |
| *From general NHS care* | 0.38137 | 0.0614 | 6.061 | 0.381370.06146.061 |
| *From non-NHS facility* | 0.5382 | 0.14563 | 3.909 | 0.53820.145633.909 |
| *Age 22 to 24* | -0.05488 | 0.41658 | -0.754 | -0.054880.41658-0.754 |
| *Age 25 to 27* | -0.32933 | 0.07201 | -4.211 | -0.329330.07201-4.211 |
| *Age 28 to 30* | -0.79636 | 0.07743 | -8.773 | -0.796360.07743-8.773 |

**Figure A1:**
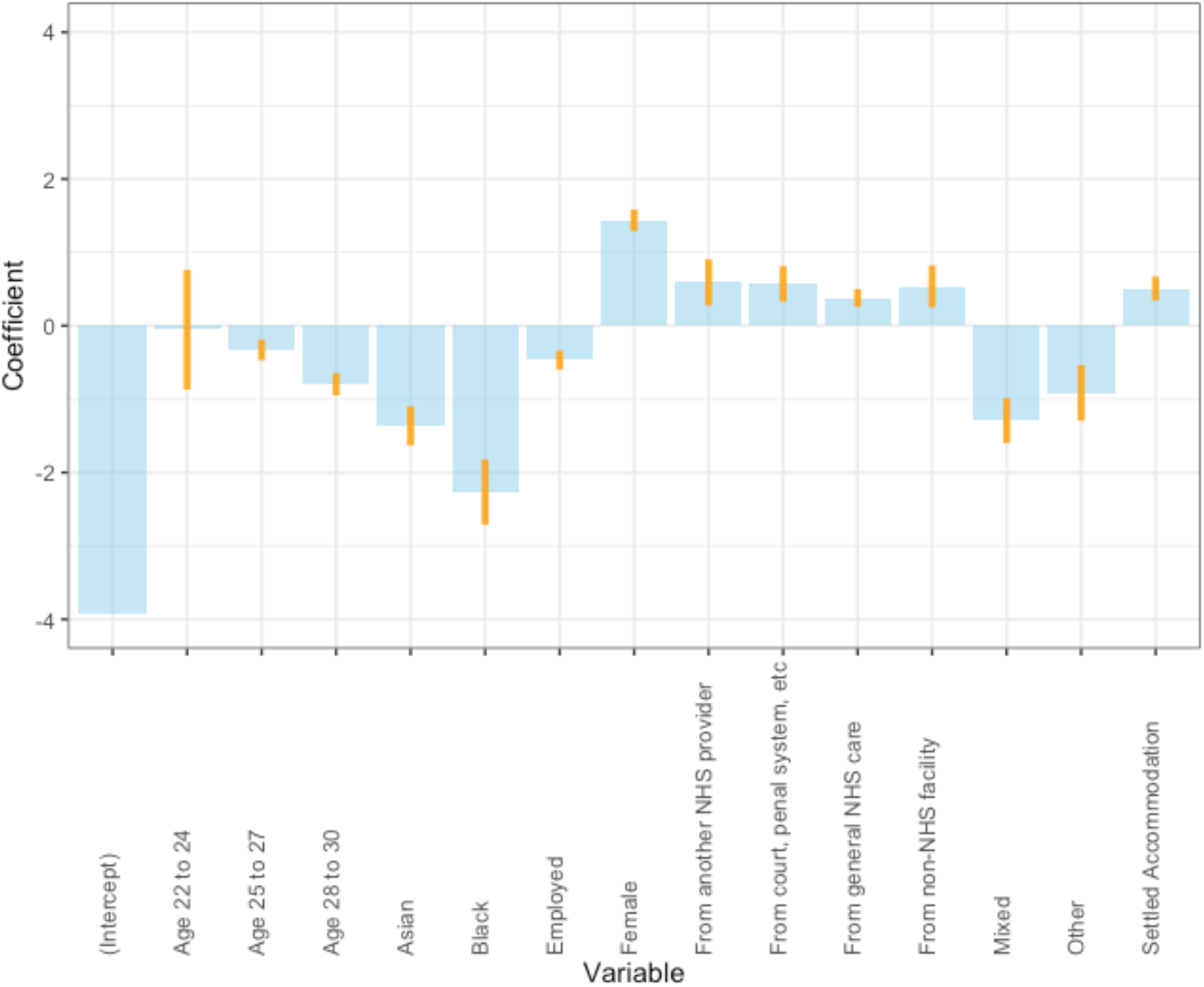
results of a generalised linear regression model to identify relationships between predictive variables and self-harm. Bars illustrate 95% confidence intervals.

